# Prevalence of screening HIV among LGBTQ+ at Ramathibodi Hospital

**DOI:** 10.1101/2025.02.21.25322658

**Authors:** Nunticha Chuenpakorn, Kewalin Chaisoksombat, Sukanya Siriyotha, Nanthiphat Chuenpakorn, Jiraporn Arunakul, Rapeephan R. Maude

**Author notes:** These authors contributed equally to this work. These authors also contributed equally to this work (Kewalin Chaisoksombat, Nanthiphat Chuenpakorn).

## Abstract

**Background:** LGBTQ+ individuals face significant barriers in accessing healthcare services, particularly in relation to HIV screening. This study investigates the prevalence of HIV testing among LGBTQ+ individuals and explores factors influencing healthcare-seeking behaviors at Ramathibodi Hospital in Bangkok, Thailand.

**Methods:** A cross-sectional survey was conducted among 300 self-identified LGBTQ+ individuals aged 18 and older who recently utilized outpatient services at Ramathibodi Hospital. Data were collected via an online questionnaire assessing demographic characteristics, sexual behaviors, and attitudes toward public health services. Univariate and multivariate logistic regression analyses were performed to identify factors associated with HIV testing.

**Results:** HIV testing prevalence among participants was 45%, with significant associations found between HIV testing and gender identity, age, and engagement in sexual activity. Gay participants (OR = 21.73, 95% CI: 4.73–99.90, p < 0.001) and transgender females (OR = 7.51, 95% CI: 2.08–27.34, p = 0.002) were more likely to undergo HIV testing compared to other groups. Those aged 30 years or older (OR = 2.50, 95% CI: 1.43– 4.34, p = 0.001) and those engaging in sexual activity (OR = 4.58, 95% CI: 2.52–8.33, p < 0.001) were also more likely to be tested. Participants reported mixed experiences regarding the inclusivity of healthcare environments, with a desire for improved LGBTQ+ cultural competence among healthcare workers.

**Conclusion:** While HIV testing rates are relatively high among certain LGBTQ+ subgroups, significant gaps remain, particularly for transgender males. The study highlights the need for targeted outreach and healthcare interventions to increase HIV screening among underserved populations. Expanding LGBTQ+ cultural competency in healthcare settings is crucial to ensure inclusive and respectful care.

## Introduction

Over the past few decades, there has been a growing recognition that gender identity extends beyond the traditional binary classification of male and female based on biological characteristics. Contemporary discussions now encompass a spectrum of identities, including cisgender, transgender men, transgender women, and nonbinary individuals ^(1, 2)^, all of whom are often collectively referred to as the LGBTQ+ community. Despite this broader understanding of gender, LGBTQ+ individuals continue to face significant barriers in accessing healthcare services. These barriers are often the result of societal norms, discrimination, and systemic inequalities, which contribute to health disparities within this population. One critical area of concern is the intersection of LGBTQ+ identities and sexually transmitted disease (STD) screening, where significant gaps in service provision remain. Research has highlighted the inadequate rates of screening for sexually transmitted infections (STIs), particularly among transgender and nonbinary (TNB) individuals, despite their elevated risk of infection.

For instance, LGBTQ+ populations, particularly gay and bisexual men, experience disproportionately high rates of HIV infection. Recent studies have shown that 44% of syphilis cases among LGBTQ+ individuals co-occur with HIV infection^(3)^. Transgender individuals are also at elevated risk, with community-based programs reporting a 2.8% HIV diagnosis rate among transgender individuals who have been tested^(4)^. The stigma surrounding both HIV and LGBTQ+ identities exacerbates these disparities by contributing to delayed testing and treatment^(5, 6)^. Furthermore, while 80.5% of TNB participants in one study reported having undergone an STI test at some point in their lives, only 59.8% had been tested within the previous year^(7)^, indicating a gap in regular screening.

The Centers for Disease Control and Prevention (CDC) recommends annual HIV screening for men who have sex with men (MSM), with more frequent testing for individuals at higher risk^(8)^. Pre-exposure prophylaxis (PrEP) has also been identified as a critical tool for preventing HIV transmission in high-risk LGBTQ+ populations, as emphasized in a recent systematic review^(9)^. However, screening for other health conditions remains inconsistent, with studies showing that LGBTQ+ individuals are screened less frequently for diseases such as cervical cancer when compared to their heterosexual counterparts^(10)^.

One of the key contributors to these disparities is the lack of adequate training among healthcare providers, who may not be fully equipped to address the unique healthcare needs of LGBTQ+ patients^(11)^. Improving the inclusivity and competence of healthcare environments is essential to ensuring that LGBTQ+ individuals receive appropriate and timely screenings, as well as broader medical care.

This study seeks to investigate the prevalence of HIV testing among LGBTQ+ individuals and explore the factors influencing their healthcare-seeking behaviors. Additionally, it aims to examine LGBTQ+ individuals’ preferences regarding healthcare services, particularly those related to inclusive environments and gender-affirming care. By identifying gaps in service provision and understanding the desires of LGBTQ+ patients, this research aims to provide valuable insights that will inform efforts to improve healthcare systems, ensuring that they are more equitable and accessible to all members of the LGBTQ+ community.

## Methodology

### Study Design

The study employs a cross-sectional survey design to examine the prevalence of HIV screening among LGBTQ+ individuals. Data acquisition is facilitated through an online questionnaire administered via Google Forms. The study utilizes a convenience sampling approach to recruit participants who have accessed health services at the outpatient clinic of Ramathibodi Hospital.

### Participants

The study population comprises individuals aged 18 years or older who self-identify as LGBTQ+ and have recently utilized health services at the outpatient clinic of Ramathibodi Hospital. A target sample size of 300 participants will be recruited. Inclusion criteria are as follows: (1) age ≥ 18 years, (2) self-identification as LGBTQ+, and (3) recent utilization of health services at the specified outpatient clinic.

### Data Collection Instrument

The primary data collection instrument is a researcher-designed online questionnaire disseminated via Google Forms. The questionnaire is structured into four distinct sections:

1. Baseline Characteristics: This section elicits demographic data, including age, gender identity, sexual orientation, and socioeconomic status.
2. Sexual Behavior: This component investigates sexual practices, number of sexual partners, and utilization of protective measures.
3. Screening for Sexually Transmitted Diseases (STDs): This section assesses participants’ history of STD testing, frequency of HIV screening, and others STD such as hepatitis B and hepatitis C etc.
4. Attitudes toward Public Health Services: This segment evaluates perceptions regarding the accessibility, quality, and efficacy of public health services related to HIV screening and sexual health.

The questionnaire underwent refinement following a pilot study involving 10-15 participants. Feedback from this pilot informed revisions to enhance the clarity, relevance, and reliability of the questions.

### Data Collection Procedure

Participants will access the online questionnaire via a distributed link. To ensure confidentiality, the questionnaire will be administered anonymously. Prior to commencing the survey, participants will be provided with comprehensive information about the study, including its purpose and the voluntary nature of their participation. Data were collected between June 2023 and December 2024. Verbal consents were documented in the electronic form from all participants and witnessed by investigators who served as independent observers to verify the voluntary participation of each respondent.

### Data Security

The Google Forms platform will be utilized for data collection. Survey responses will be securely stored, with access restricted solely to the principal investigator. Stringent measures will be implemented to ensure data privacy and integrity, including password protection and restricted access protocols.

### Data Analysis

Descriptive statistics will be employed to summarize the demographic characteristics, sexual behaviors, history of sexually transmitted disease (STD) screening, and attitudes toward public health services within the study population. Continuous variables will be presented as mean ± standard deviation (SD) or median (range), while categorical variables will be described using frequencies and percentages. Comparisons between participants who have undergone HIV screening and those who have not will be made using the Student’s t-test or Wilcoxon rank-sum test for continuous variables, and the Chi-square test or Fisher’s exact test for categorical variables.

To identify factors associated with HIV screening, both univariate and multivariate logistic regression analyses will be conducted. Variables with a p-value of less than 0.1 in the univariate analysis will be included in the multivariate model. Only variables that remain significant according to the likelihood ratio test will be retained in the final model. All statistical analyses will be performed using STATA version 18.0 (StataCorp LLC, College Station, TX, USA), and a p-value of less than 0.05 will be considered statistically significant.

### Ethical Considerations

This study will adhere to established ethical standards in research, including obtaining electronic informed consent from all participants. The study protocol will undergo review and approval by the Institutional Review Board (IRB) of Ramathibodi Hospital (COA.MURA2023/440). Data confidentiality will be maintained, and all identifying information will be anonymized to protect participant privacy.

## Results and Discussion

The study included 300 participants, of whom 29.67% were biological male at birth and 70.33% were biological female at birth. In terms of gender identity, the largest group was transgender males (55.67%), followed by transgender females (15.67%), queer individuals (10%), and gay individuals (9.33%). The median gender transition duration was 45 (IQR 17,115) months, with 50.67% of participants reporting a transition of 45 months or longer. The mean age of participants was 32.16 ± 8.28 years, and 57% were aged 30 years or older. Regarding education, the majority held a bachelor’s degree (71.33%), while 14% had a postgraduate degree, and 14.67% had completed elementary or high school. The most common occupations were office workers (43.33%), business owners or freelancers (22.67%), and government officers (17.67%). The median monthly salary was 25,000 (IQR 18000,40000) THB, with 53.67% of participants earning 25,000 THB or more per month (Table 1).

**Table 1.**
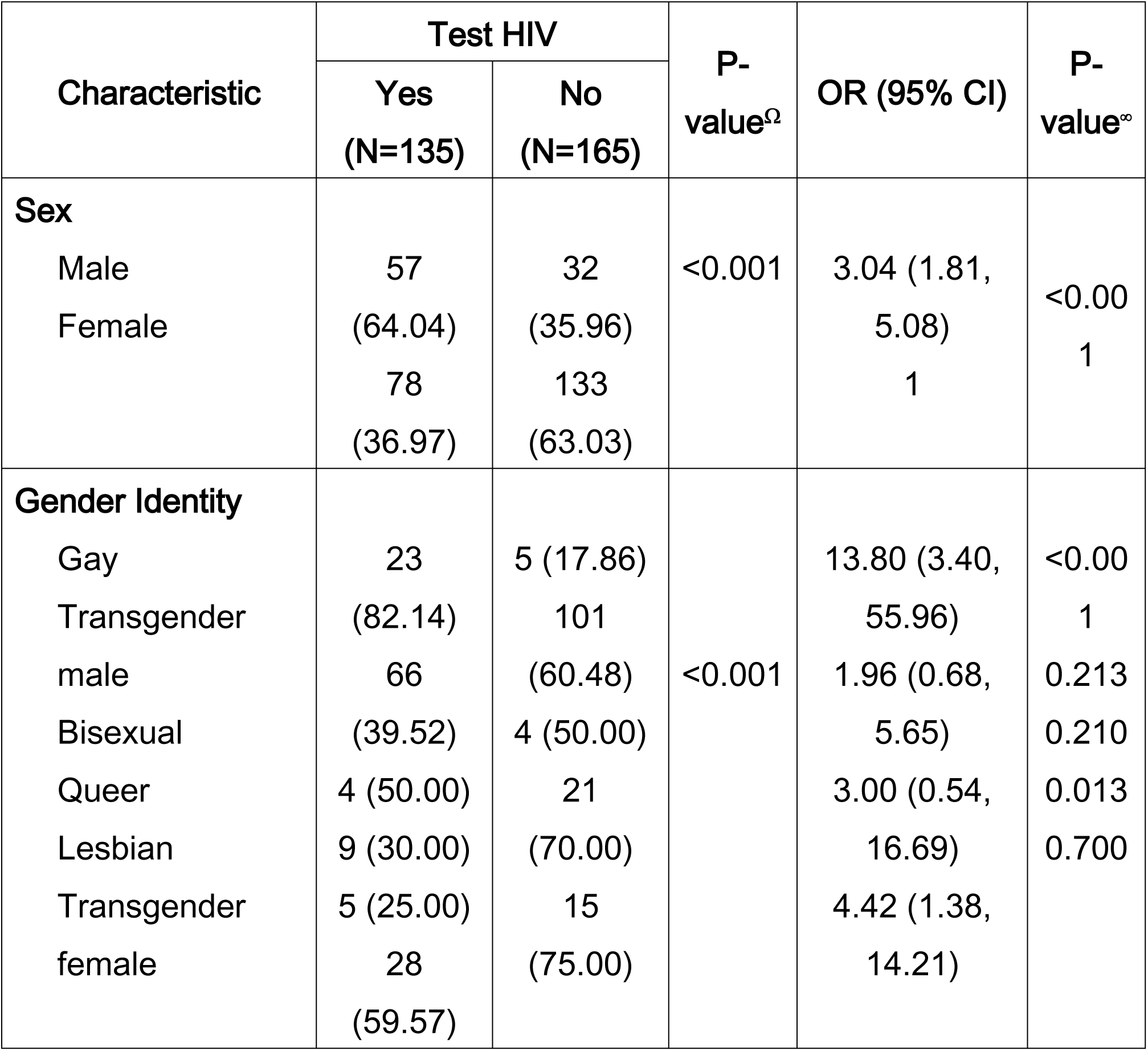
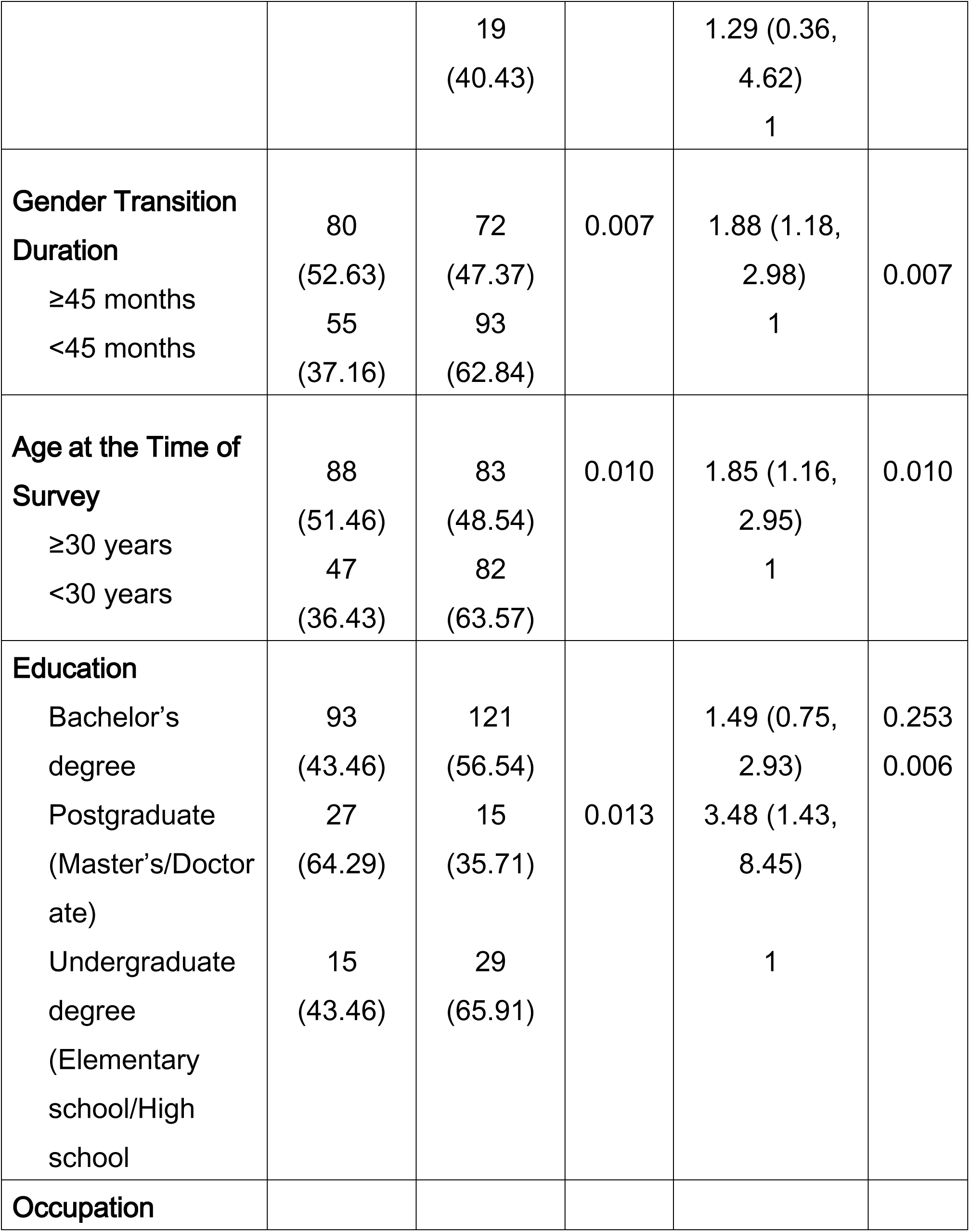
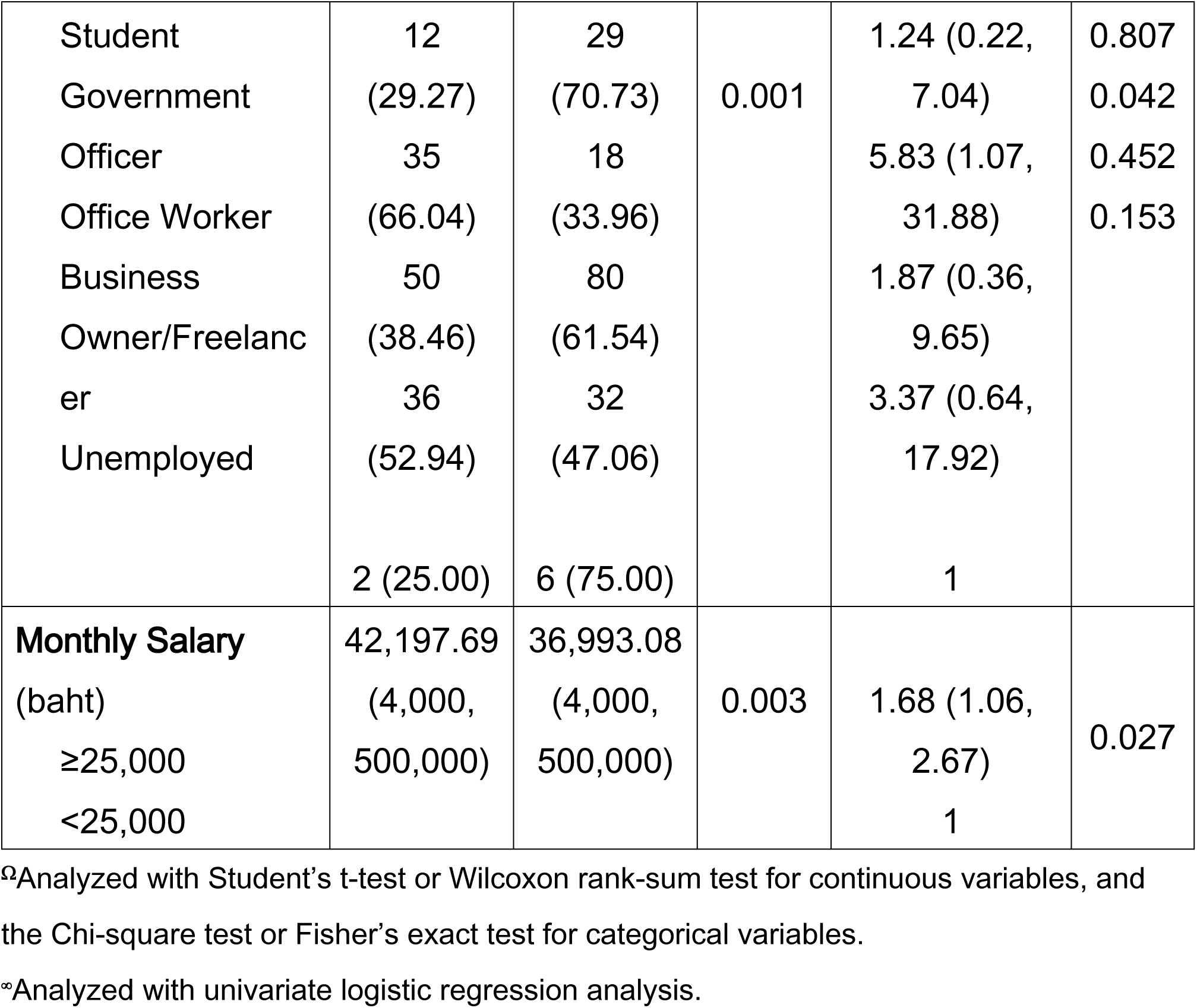
Participants Characteristics Associated with HIV Testing.

In terms of sexual behavior, 68% of participants reported having engaged in sexual activity, with a median of 4 (IQR 2,6) sexual partners. The most common methods of sexual engagement were oral sex (70.59%) and penetrative sex (68.14%). Regarding the use of protection during sexual activity, 60.78% of participants reported using protection, with condoms being the most common method (97.58%) (Table 2). A total of 45% of participants had undergone HIV testing, with 77.78% of these individuals being tested at a hospital. However, other sexually transmitted infections (STIs) such as hepatitis B (9.67%), hepatitis C (4.33%), gonorrhea (1.33%), and chlamydia (0.67%) were tested at much lower rates (Table 3). Participants also expressed a desire for their gender identity to be included in their medical records (74.33%), and 83.33% reported finding healthcare facilities such as toilets convenient.

**Table 2.**
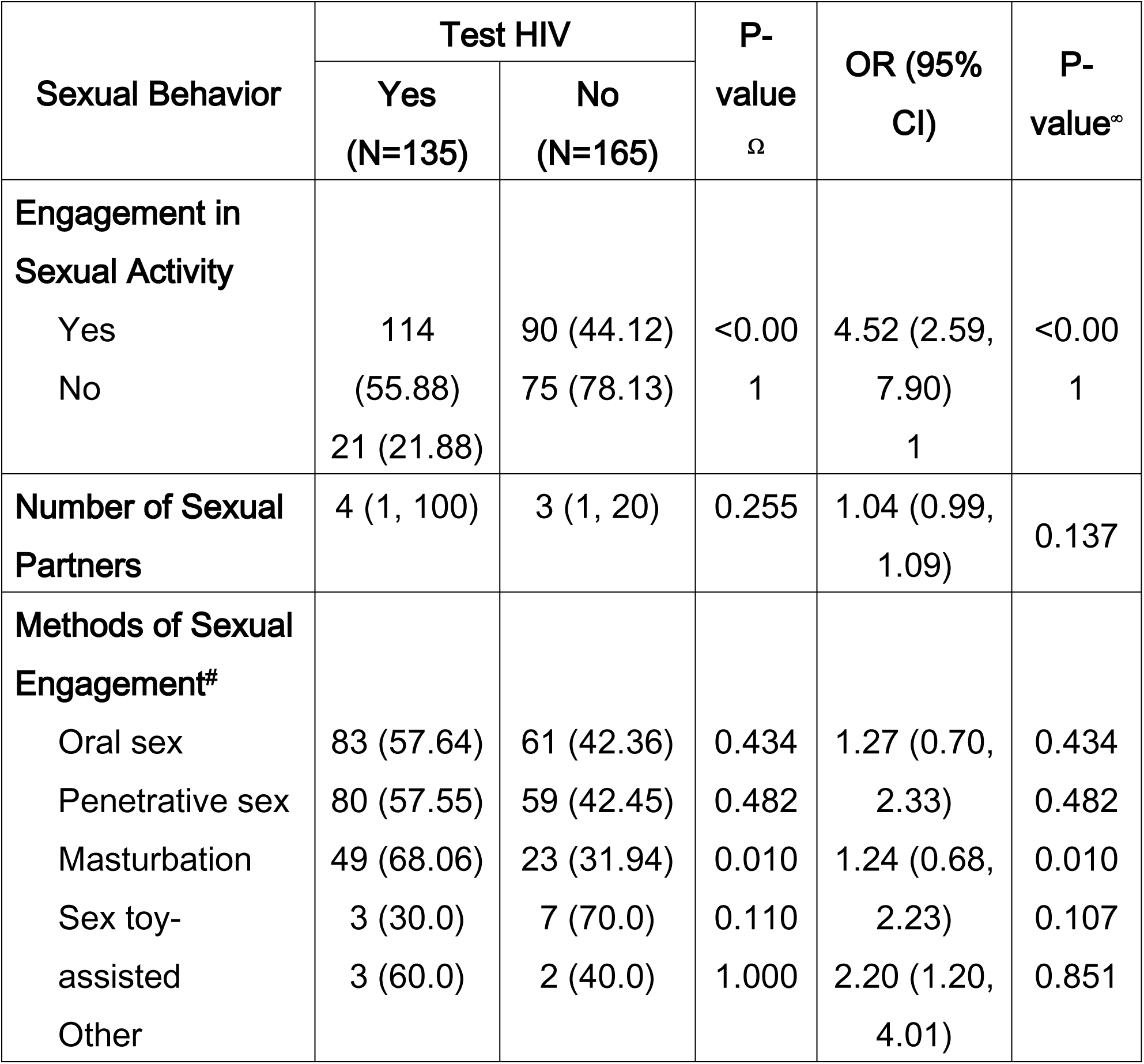
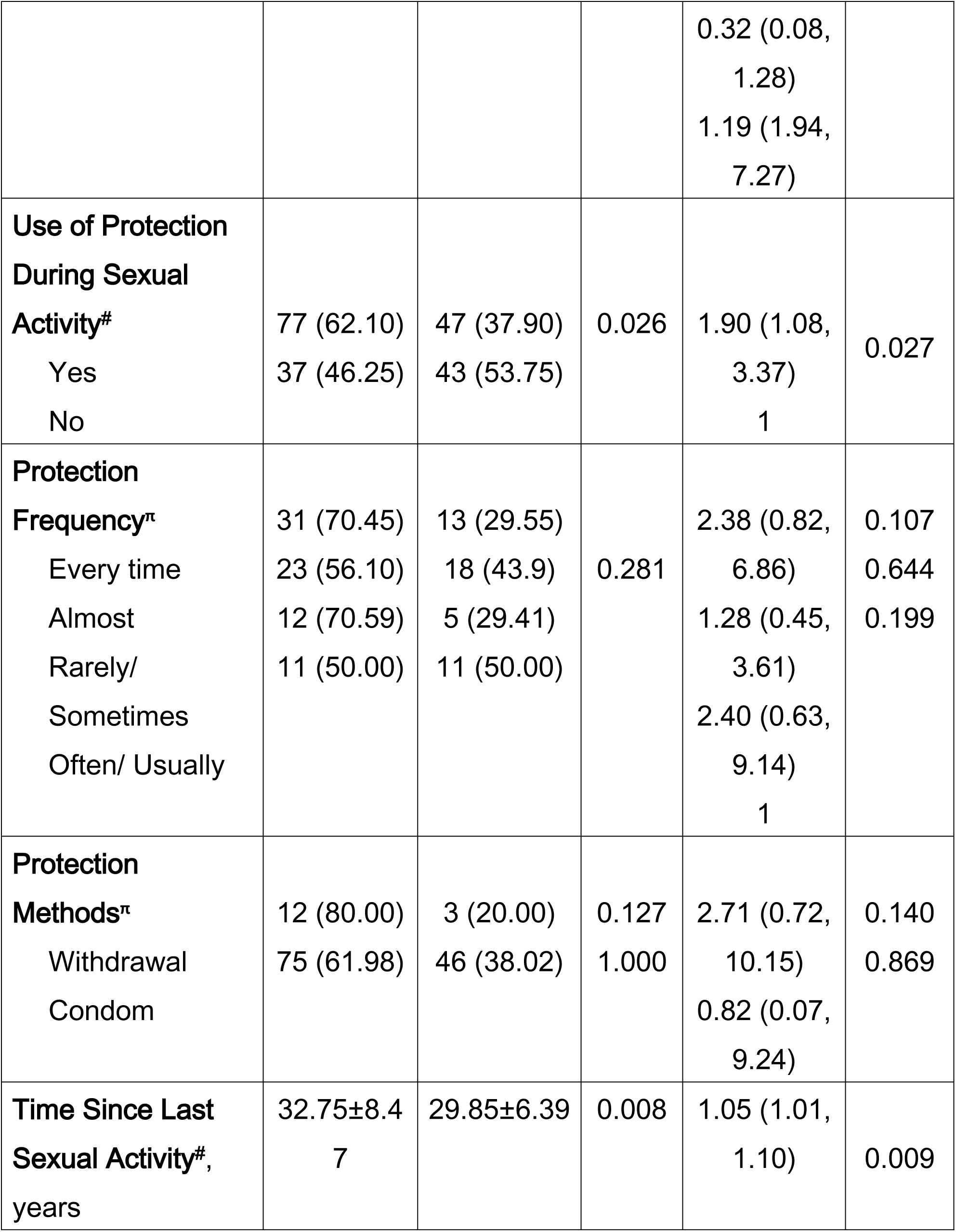
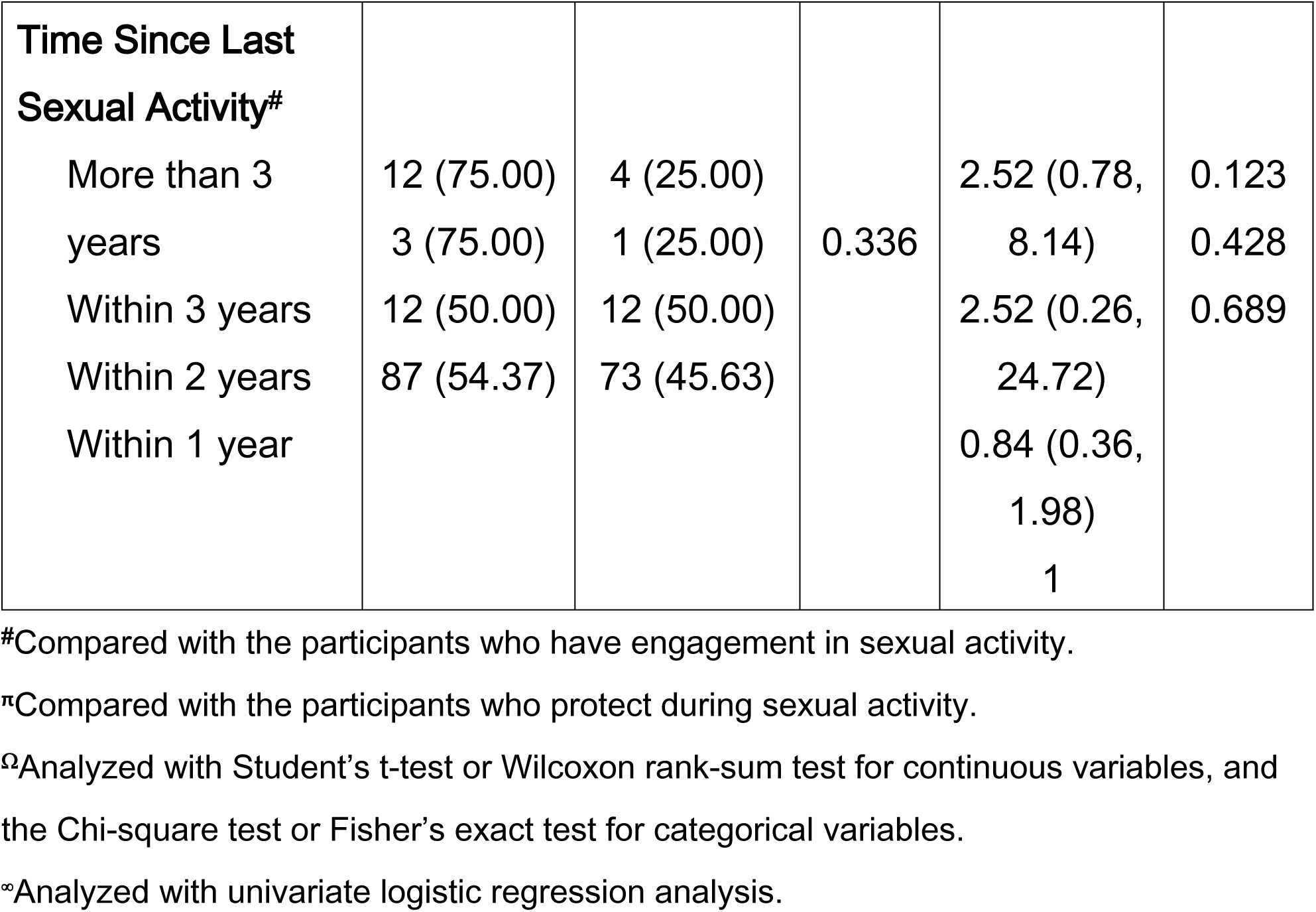
Sexual Behavior Associated with HIV Testing.

**Table 3.**
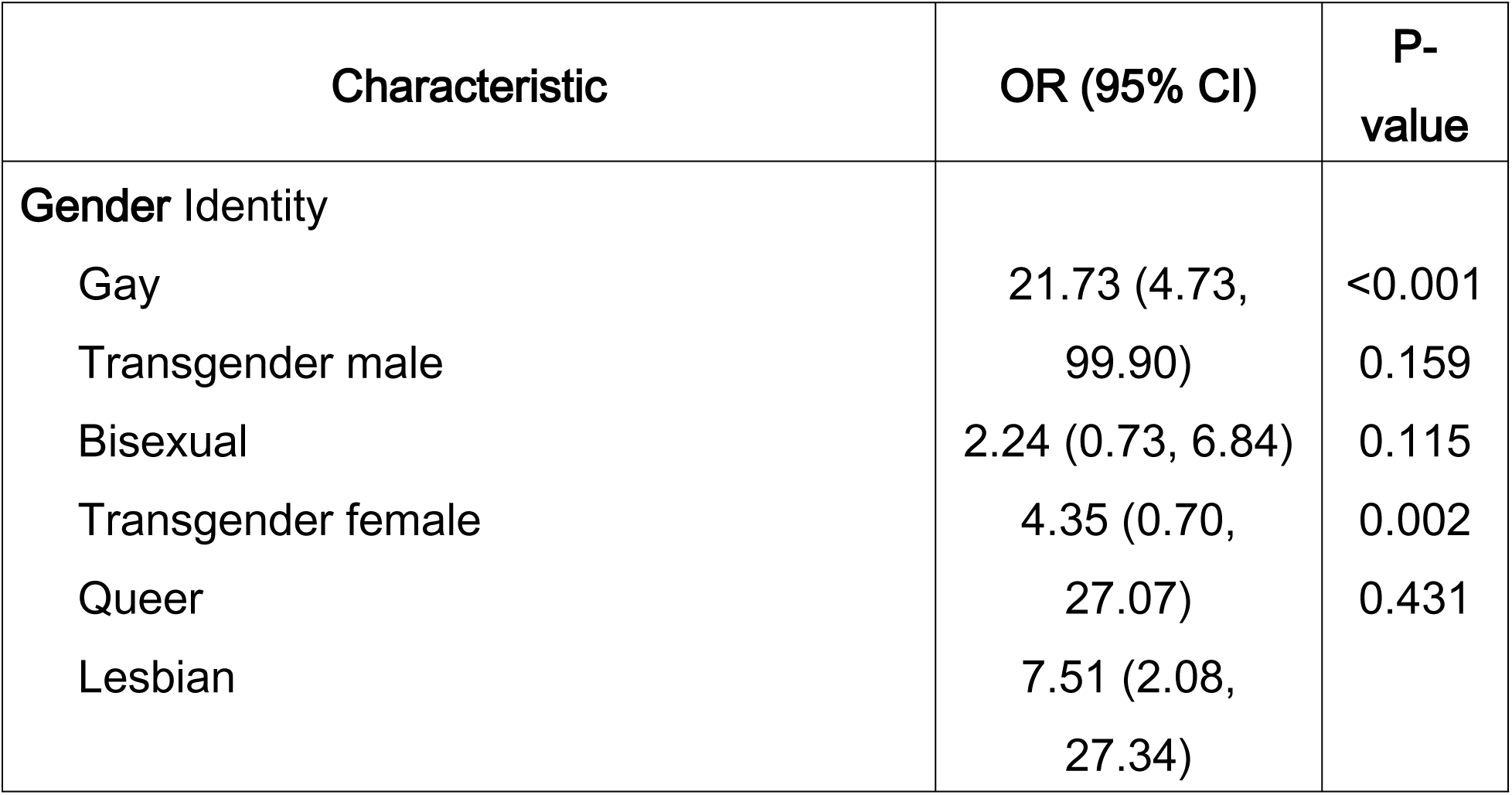
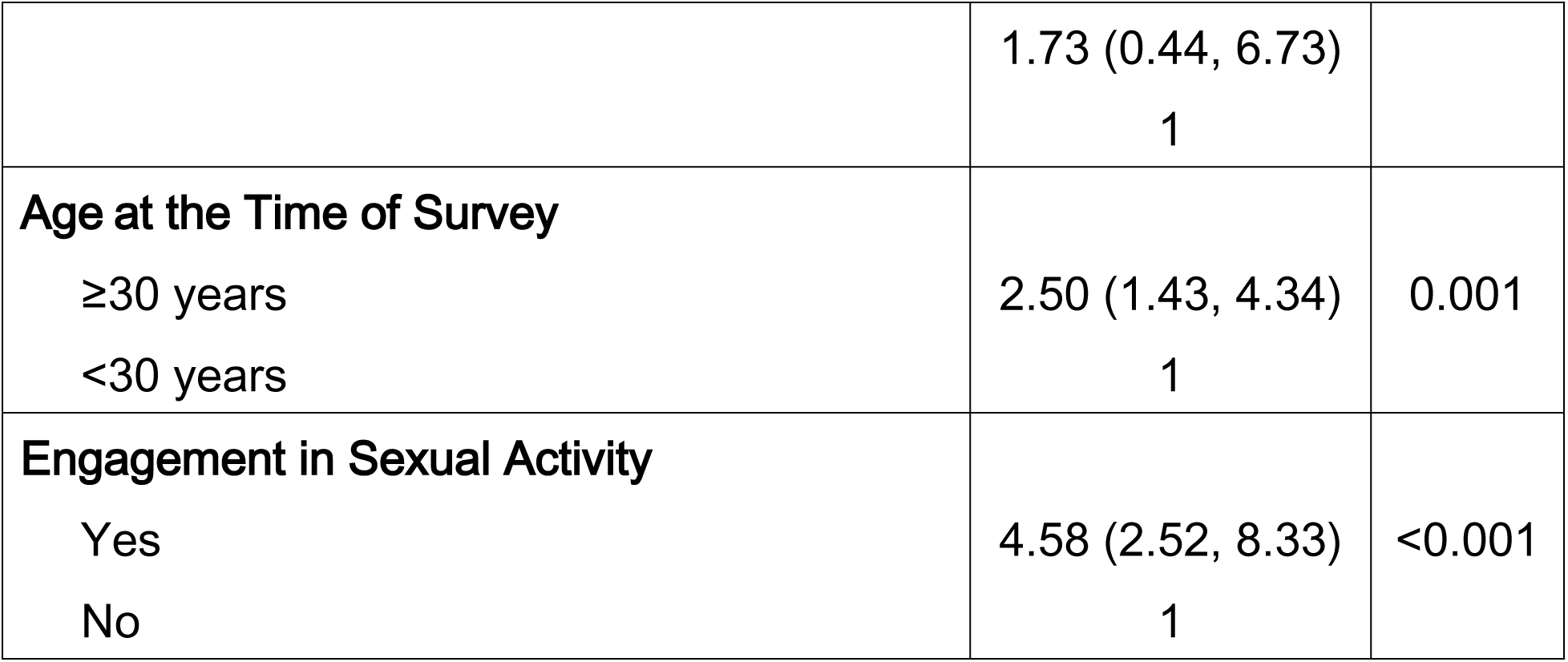
Multivariate Analysis of Factors Associated with HIV Testing.

With regard to medical preferences, 74.33% of participants expressed a desire for the inclusion of their gender identity in medical records, and 37.67% preferred to use an alias. For service calls, 63% favored their real name, 39.33% preferred their nickname, and 20.67% opted for an alias. Most participants (83.33%) reported comfort in using available restroom facilities. However, the perceived understanding of healthcare workers toward LGBTQ+ individuals received a mean score of 3.66 ± 1.15, reflecting mixed experiences (Table 4). Some participants felt that healthcare professionals lacked knowledge of LGBTQ+ identities, with transgender individuals particularly highlighting discomfort when treated according to their assigned gender at birth rather than their gender identity. Additionally, some staff continued to use disrespectful language or displayed behaviors that were perceived as discriminatory. The lack of exposure to LGBTQ+ issues in certain regions and the healthcare system, especially in smaller provinces, was also noted. On the other hand, some participants reported positive experiences where healthcare workers were respectful and used preferred names, showing that understanding varied widely across healthcare settings. These inconsistencies underline the need for better education and training in gender diversity within the healthcare system.

**Table 4.** Medical Preferences.

| **Preferences** | **N (%)** |
| --- | --- |
| **Inclusion of Gender Identity in Medical Records** |  |
| Yes | 223 (74.33) |
| No | 77 (25.67) |
| **Preference for Alias in Medical Records** |  |
| Yes | 113 (37.67) |
| No | 187 (62.33) |
| **Preferred Name for Service call** |  |
| Real Name | 189 (63.00) |
| Nickname | 118 (39.33) |
| Alias | 62 (20.67) |
| Queue Number | 4 (1.33) |
| Prefix | 3 (1.00) |
| No Preference | 4 (1.33) |
| **Convenience in Using Toilets** |  |
| Yes | 250 (83.33) |
| No | 50 (16.67) |
| **Score Perception of Healthcare Workers' Understanding of LGBTQ+, median±SD** | 3.66±1.15 |

In terms of health screenings, 28% reported having been screened for diabetes, with 54.76% screened before the recommended age of 35 years. Dyslipidemia screening was more common, with 52.33% undergoing tests, and 62.22% of these tests being conducted at the recommended age (≥35 years). Cervical cancer screening was reported by 34.12% of biological females, with a mean screening age of 34.99 ± 7.47 years. Additionally, 22.33% of participants received the HPV vaccine, though only 24.00% were vaccinated within the recommended age range (9-45 years). Colon cancer screening was reported by 6.33% of participants, with only 8% screened at the proper age of 45 years or older (Table 5).

**Table 5.** Screening and Health Services.

| Screening | N (%) |
| --- | --- |
| **Diabetes Screening** |  |
| Yes | 84 (28.00) |
| No | 216 (72.00) |
| **Proper Age Screening<sup>a</sup> (n=90)** |  |
| Yes | 38 (42.22) |
| No | 52 (57.78) |
| **Dyslipidemia Screening** |  |
| Yes | 157 (52.33) |
| No | 143 (47.67) |
| **Proper Age Screening<sup>α</sup> (n=90)** |  |
| Yes | 56 (62.22) |
| No | 34 (37.78) |
| **Cervical Cancer Screening (for biological female)** |  |
| Yes | 72 (34.12) |
| No | 139 (65.88) |
| **Received HPV Vaccination** |  |
| Yes | 67 (22.33) |
| No | 233 (77.67) |
| **Colon Cancer Screening** |  |
| Yes | 19 (6.33) |
| No | 281 (9.367) |
| **Proper Age Screening<sup>β</sup> (n=25)** |  |
| Yes | 2 (8.00) |
| No | 23 (92.00) |

<sup>α</sup>Screening age at 35 years and older
<sup>β</sup>Screening age at 45 years and older

The univariate analysis revealed that several factors were significantly associated with HIV testing. Male participants were more likely to have undergone HIV testing than female participants (OR = 3.04, 95% CI: 1.81–5.08, p < 0.001). Gay participants were significantly more likely to have been tested for HIV compared to other gender identities (OR = 13.80, 95% CI: 3.40–55.96, p < 0.001), followed by transgender females (OR = 4.42, 95% CI: 1.38–14.21, p = 0.013). A longer gender transition duration (≥45 months) was also associated with a higher likelihood of HIV testing (OR = 1.88, 95% CI: 1.18–2.98, p = 0.007). Age was another significant factor, with those aged 30 years or older being more likely to have undergone HIV testing (OR = 1.85, 95% CI: 1.16–2.95, p = 0.010). Educational attainment was also significant, with participants holding postgraduate degrees more likely to have undergone HIV testing (OR = 3.48, 95% CI: 1.43–8.45, p = 0.006) (Table 1).

Similarly, the univariate analysis of sexual behavior-related factors identified significant associations with HIV testing. Participants who reported engaging in sexual activity were significantly more likely to have been tested for HIV compared to those who had not (OR = 4.52, 95% CI: 2.59–7.90, p < 0.001). The use of protection during sexual activity was also associated with higher rates of HIV testing (OR = 1.90, 95% CI: 1.08–3.37, p = 0.027). Furthermore, the method of sexual engagement influenced testing likelihood, with individuals engaging in masturbation being more likely to have undergone HIV testing (OR = 2.20, 95% CI: 1.20–4.01, p = 0.010). Time since last sexual activity also emerged as a significant factor, with more recent sexual activity correlating with an increased likelihood of HIV testing (OR = 1.05, 95% CI: 1.01–1.10, p = 0.009) (Table 2).

Multivariate analysis confirmed that gender identity, age, and engagement in sexual activity were significant factors associated with HIV testing. Gay participants were much more likely to have been tested for HIV (OR = 21.73, 95% CI: 4.73–99.90, p < 0.001), followed by transgender females (OR = 7.51, 95% CI: 2.08–27.34, p = 0.002). Those aged 30 years or older were more likely to have been tested for HIV (OR = 2.50, 95% CI: 1.43–4.34, p = 0.001). Engagement in sexual activity remained a strong predictor, with those reporting sexual activity being 4.58 times more likely to have undergone HIV testing (OR = 4.58, 95% CI: 2.52–8.33, p < 0.001) (Table 3).

## Discussion

This study provides valuable insights into the healthcare experiences of LGBTQ+ individuals, particularly in the areas of HIV testing, sexual health services, and the perceived understanding of healthcare workers regarding gender and sexual diversity. Consistent with previous research, the findings reveal that while a significant portion of LGBTQ+ individuals, particularly gay and transgender females, engage in HIV testing, there remain substantial gaps in service access, particularly among transgender males^(12, 13)^. These disparities highlight the continued need for tailored outreach efforts and healthcare interventions to ensure that all subgroups, particularly those less engaged in regular testing, receive appropriate and inclusive care^(14)^.

The high prevalence of HIV testing among gay and transgender females aligns with targeted public health campaigns that have successfully reached these populations^(12)^. However, the lower testing rates among transgender males reflect a concerning gap in outreach and services specifically tailored to their needs, as highlighted in studies on STI prevention efforts^(13)^. This gap may also be influenced by socioeconomic factors, such as access to healthcare and education, as research shows that LGBTQ+ individuals, particularly transgender people, often face economic inequalities and health inequities^(15)^. Individuals with longer gender transition durations and those aged 30 years or older were more likely to undergo HIV testing, possibly indicating increased health awareness in these populations. This finding underscores the importance of developing targeted health education strategies that reach younger LGBTQ+ individuals or those earlier in their gender transition journeys^(14, 16)^.

Moreover, the study found that sexual behavior, including masturbation, significantly influenced HIV testing rates. This suggests that individuals with more varied sexual practices may perceive themselves at higher risk of infection, prompting them to seek testing^(17)^. Additionally, recent sexual activity was another key predictor of testing, with more frequent engagement correlating with higher testing rates. These findings are consistent with literature indicating that sexual activity serves as a driver for healthcare-seeking behavior, particularly for STI screening^(18)^. Such behaviors further underscore the need for comprehensive sexual health services that cater to a wide range of practices and are inclusive of diverse LGBTQ+ identities.

A key area of concern raised by the study is the inconsistent understanding of LGBTQ+ identities among healthcare workers. Many participants reported discriminatory practices or a lack of respect for their gender identity, such as being addressed by their birth-assigned gender or encountering inappropriate language. These experiences were more common in smaller healthcare facilities or rural areas, which may lack exposure to LGBTQ+ health issues^(19)^. This is consistent with previous research that identifies gaps in healthcare worker education on LGBTQ+ health^(12)^. Improving the training and cultural competence of healthcare professionals is crucial to fostering more inclusive and respectful healthcare environments. By ensuring that healthcare workers are equipped with the knowledge and sensitivity to address LGBTQ+ patients, we can improve the quality of care and reduce disparities in health outcomes^(14)^.

Furthermore, the study’s findings indicate a strong preference among participants for having their gender identity included in medical records and being addressed by their preferred names. This reflects broader demands within the LGBTQ+ community for greater recognition and respect of their gender identity in healthcare settings^(16)^. Adopting policies that support more flexible gender identification and training healthcare professionals to address patients appropriately can significantly enhance patient comfort, trust, and engagement with healthcare systems.

Despite significant progress in promoting HIV testing among LGBTQ+ populations, the study also highlights the continued disparities in the screening of other sexually transmitted infections (STIs), such as hepatitis B, hepatitis C, gonorrhea, and chlamydia. These low testing rates suggest that comprehensive sexual health services may not be as readily available or promoted, particularly for transgender individuals and those in rural or underserved areas^(12, 13)^. Addressing these disparities requires expanding access to a broader range of sexual health services, ensuring that LGBTQ+ individuals receive the full spectrum of care, including STI screening beyond HIV^(14)^.

Looking toward the future, technological advancements present a promising avenue for increasing awareness, care, and the uptake of sexual health services among LGBTQ+ populations. Digital platforms and online interventions, such as web-based dramas or mobile health applications, have shown promise in promoting HIV and STI testing, particularly among younger gay and bisexual men^(17)^. Moreover, studies suggest that technology can be harnessed to deliver targeted health messaging and support systems to sexual and gender minority adults in underserved areas, including those in East Africa^(20)^. Expanding the use of digital tools, including telemedicine and mobile health initiatives, could help bridge the gaps in healthcare access, particularly for those in rural areas or with limited access to LGBTQ+ friendly healthcare facilities. By integrating technology into public health campaigns and healthcare delivery, we may be able to enhance outreach efforts and ensure more equitable and inclusive access to healthcare services across the LGBTQ+ community.

## Limitations and Recommendations

This study was conducted at a single institution in Bangkok, the capital city of Thailand, where the educational level and background knowledge of the population may differ from other regions of the country. Consequently, the findings may not fully reflect the experiences of LGBTQ+ individuals in different healthcare settings across Thailand. Additionally, the reliance on self-reported data introduces the possibility of social desirability bias, and the use of an online questionnaire may have excluded individuals without internet access or those less comfortable with digital formats. Future research should aim to conduct larger, multicenter studies to provide a more comprehensive understanding of HIV screening behaviors among LGBTQ+ populations across different regions of Thailand. Expanding the scope to include larger online questionnaires and in-depth interviews would also offer deeper insights into preferences and experiences regarding medical services, particularly in terms of inclusivity and accessibility.

## Conclusion

This study sheds light on the experiences of LGBTQ+ individuals within healthcare systems, emphasizing the need for more inclusive and respectful care. The variability in HIV testing rates among different subgroups suggests that targeted outreach and education are necessary to ensure that all LGBTQ+ individuals receive appropriate sexual health services. Furthermore, the inconsistencies in healthcare workers’ understanding of LGBTQ+ identities highlight the ongoing need for comprehensive training programs aimed at improving the cultural competence of healthcare providers. By addressing these gaps, healthcare systems can improve the quality of care for LGBTQ+ individuals, fostering a more inclusive environment that respects their identities and meets their specific health needs.

In conclusion, while progress has been made in recognizing the healthcare needs of LGBTQ+ populations, significant work remains to be done. Expanding healthcare workers’ knowledge of gender diversity, increasing access to comprehensive sexual health services, and ensuring that LGBTQ+ individuals feel respected and understood in healthcare settings are critical steps toward achieving health equity for this population.

## Data Availability

All relevant data are within the manuscript and its Supporting Information files.

## Acknowledgments

This research project is supported by Faculty of Medicine Ramathibodi Hospital, Mahidol University (RF_66102)

Kewalin Chaisoksombat^3&^, Nanthiphat Chuenpakorn^&^, Teeraporn Janchai^1&^, Sukanya Siriyotha^4&^

## Acknowledgments

We would like to express our sincere gratitude to all patients who utilized the healthcare services at GenV Clinic, Ramathibodi Hospital, for their participation in this study. This research was supported by funding from the Faculty of Medicine, Ramathibodi Hospital (RF_66102). Special thanks are due to the outpatient nurse and research assistant who actively supported the recruitment and advertisement efforts. We extend our deep appreciation to Nanthiphat for his invaluable assistance in data collection, and to Miss Sukanya, our statistician, for her expert guidance in data analysis. This research would not have been possible without their essential contributions.

## References

1. AEC F. Defining LGBTQ Terms and Concepts 2023.

2. Sabra L. Katz-Wise P. Gender fluidity: What it means and why support matters 2020 [cited 2023 3 March].

3. Alawiyah BHS, Mawaddah A, Indrasari AD, Lestari AR, Wahyudi D, Ahda FR, et al. Most Common Sexually Transmitted Infections in LGBT. Jurnal Biologi Tropis. 2023;23(1):62–7.

4. Mulatu MS, Wang G, Song W, Keatley J, Kudon HZ, Wan C, et al. Brief Report: HIV Testing, Diagnosis of HIV Infection, Linkage to Medical Care, and Interview for Partner Services Among Transgender Persons-United States, 2012-2017. J Acquir Immune Defic Syndr. 2021;86(5):530-5.

5. Nguyen TT, Do AL, Nguyen LH, Vu GT, Dam VAT, Latkin CA, et al. Scholarly literature in HIV-related lesbian, gay, bisexual, and transgender studies: A bibliometric analysis. Front Psychol. 2023;14:1028771.

6. Tomar A, Spadine MN, Graves-Boswell T, Wigfall LT. COVID-19 among LGBTQ+ individuals living with HIV/AIDS: psycho-social challenges and care options. AIMS Public Health. 2021;8(2):303–8.

7. Lacombe-Duncan A, Kattari SK, Kattari L, Scheim AI, Misiolek BA. Sexually transmitted infection testing among transgender and non-binary persons: results of a community-based cross-sectional survey. Sex Health. 2023;20(1):87–91.

8. DiNenno EA, Prejean J, Irwin K, Delaney KP, Bowles K, Martin T, et al. Recommendations for HIV Screening of Gay, Bisexual, and Other Men Who Have Sex with Men - United States, 2017. MMWR Morb Mortal Wkly Rep. 2017;66(31):830-2.

9. Dorce-Medard DJ, Okobi Md OE, Grieb DJ, Saunders DN, Harberger Md S. HIV Pre-exposure Prophylaxis in the LGBTQ Community: A Review of Practice and Places. Cureus. 2021;13(6):e15518.

10. Grasso C, Goldhammer H, Brown RJ, Furness BW. Using sexual orientation and gender identity data in electronic health records to assess for disparities in preventive health screening services. Int J Med Inform. 2020;142:104245.

11. Nelson NG, Lombardo JF, Shimada A, Ruggiero ML, Smith AP, Ko K, et al. Physician Perceptions on Cancer Screening for LGBTQ+ Patients. Cancers (Basel). 2023;15(11).

12. Burgess S, Beltrami J, Kearns L, Gruber D. The Louisiana Wellness Centers Program for HIV/STD Prevention Among Gay and Bisexual Men and Transgender Persons. J Public Health Manag Pract. 2020;26(6):590–4.

13. Pitasi MA, Kerani RP, Kohn R, Murphy RD, Pathela P, Schumacher CM, et al. Chlamydia, Gonorrhea, and Human Immunodeficiency Virus Infection Among Transgender Women and Transgender Men Attending Clinics that Provide Sexually Transmitted Disease Services in Six US Cities: Results From the Sexually Transmitted Disease Surveillance Network. Sex Transm Dis. 2019;46(2):112–7.

14. Neumann MS, Finlayson TJ, Pitts NL, Keatley J. Comprehensive HIV Prevention for Transgender Persons. Am J Public Health. 2017;107(2):207–12.

15. Folayan MO, Yakusik A, Enemo A, Sunday A, Muhammad A, Nyako HY, et al. Socioeconomic inequality, health inequity and well-being of transgender people during the COVID-19 pandemic in Nigeria. BMC Public Health. 2023;23(1):1539.

16. Mora M, Rincon G, Bourrelly M, Maradan G, Freire Maresca A, Michard F, et al. Living conditions, HIV and gender affirmation care pathways of transgender people living with HIV in France: a nationwide, comprehensive, cross-sectional, community-based research protocol (ANRS Trans&HIV). BMJ Open. 2021;11(12):e052691.

17. Tan RKJ, Koh WL, Le D, Banerjee S, Chio MT, Chan RKW, et al. Effect of a Popular Web Drama Video Series on HIV and Other Sexually Transmitted Infection Testing Among Gay, Bisexual, and Other Men Who Have Sex With Men in Singapore: Community-Based, Pragmatic, Randomized Controlled Trial. J Med Internet Res. 2022;24(5):e31401.

18. Guerra FM, Salway TJ, Beckett R, Friedman L, Buchan SA. Review of sexualized drug use associated with sexually transmitted and blood-borne infections in gay, bisexual and other men who have sex with men. Drug Alcohol Depend. 2020;216:108237.

19. Mwaniki SW, Kaberia PM, Mugo PM, Palanee-Phillips T. "What if I get sick, where shall I go?": a qualitative investigation of healthcare engagement among young gay and bisexual men in Nairobi, Kenya. BMC Public Health. 2024;24(1):52.

20. Ybarra M, Nyemara N, Mugisha F, Garofalo R. Opportunities to Harness Technology to Deliver HIV Prevention / Healthy Sexuality Programming to Sexual and Gender Minority Adults Living in East Africa. AIDS Behav. 2021;25(4):1120–8.

